# B-cell involvement supports anti-CD20 therapy in paediatric rheumatic heart disease: Proof of concept and initial safety

**DOI:** 10.64898/2026.11.17.26367998

**Authors:** Xavier Jouven, Magalie Kaya Samba, Hanifa Ismael, Halifa Farchati, Alexandre Loupy, Olivier Hermine, Marie Antignac, Patrick Bruneval, Wolf Herman Fridman, Hafid Ait Oufella, Lamine Sy, Elisabeth Tine, Ndeye Awa Sarr, Amadou Lamine Fall, Emma Menant, Ibrahima Bara Diop

## Abstract

Rheumatic heart disease (RHD) remains a major cause of cardiovascular morbidity and mortality, particularly in low- and middle-income countries, yet no therapy targets its underlying autoimmune mechanisms.[1],[2] We combined transcriptomic analyses of rheumatic mitral valves tissues with a translational approach. Using a panel of 750 mRNAs, we found a marked upregulation of B-cell-related pathways, including CD20-associated and multiple immunoglobulin genes, supporting a pathogenic role of B cells in RHD. We therefore evaluated an anti-CD20 B-cell depleting monoclonal antibody (Rituximab), in a feasibility study in children with RHD, in Dakar, Sénégal. Treatment was well tolerated, and in the following RCT first year preliminary safety analysis asked by the ethical committee, children who received rituximab showed a decrease in anti-streptolysin O titers and clinical improvement compared to controls. This proof-of-concept reveals B cells as a therapeutic target and suggests that an anti-CD20 agent may represent a potential disease-modifying therapy for RHD.

## Introduction

Rheumatic heart disease (RHD) remains a major global public health problem, affecting more than 40 million people worldwide and causing 300,000 to 400,000 deaths each year.[1] The burden of disease is disproportionately high in sub-Saharan Africa, South Asia, and parts of Oceania.[2] Although age-standardized mortality rates from RHD have declined since 1990, the absolute number of cases and deaths has remained high due to population growth.[3] Consequently, RHD is expected to persist as a leading cause of preventable cardiovascular morbidity and mortality, particularly among children, adolescents, and young adults in low-resource settings.[4]

RHD is an autoimmune condition triggered by untreated Streptococcus pyogenes pharyngitis. In susceptible individuals, an abnormal immune response leads to acute rheumatic fever (ARF), characterized by a range of clinical manifestations including pancarditis, Sydenham’s chorea, and arthritis.[5] Recurrent episodes of ARF are common and result in progressive inflammatory and fibrotic damage to the cardiac valves, predominantly the mitral valve. This process often leads to valvular regurgitation or stenosis, repeated episodes of heart failure, premature death in children, and chronic heart failure in survivors.[3]

Countries such as Australia, New-Zealand, Uganda and South Africa have made substantial investments in RHD, generating more epidemiological, clinical, genetic, echocardiographic, and therapeutic studies.[6],[7],[8],[9],[10],[11] However, the immune mechanisms underlying RHD pathogenesis remains incompletely understood.

Current management strategies are first directed against the bacterial infection. Primary prevention relies on oral penicillin to treat streptococcal infections, while secondary prevention consists of intramuscular benzathine penicillin administered every three weeks, often until the age of 21 years.[3] Although this prevention has proven to be effective in randomized control trials (RCT), its implementation is limited in practice due to the complications and potential risks associated with repeated intramuscular injections, including anaphylactic shock and death, leading to the suppression of its use in many countries.[12] Treatment of ARF episodes and heart failure is largely symptomatic, with valve replacement surgery available only to a minority of patients because of the limited availability of cardiac surgical infrastructure in endemic regions. Importantly, no specific therapy currently targets the underlying immune-inflammatory mechanisms of RHD, representing a major therapeutic gap.

To address this issue, we investigated RHD immunopathophysiology. We explored gene expression patterns in diseased mitral valves using advanced immunologic tools, to identify key molecular and cellular mechanisms involved in RHD. Based on these findings, we evaluated the rationale for targeting CD20-positive B-cells and report the interim safety analysis of the first year (as required by the ethical committee) of an ongoing RCT assessing the potential therapeutic role of an anti-CD20 agent (rituximab) in children with RHD.[13],[14],[15]

## Results

### Transcriptomic analysis of rheumatic mitral valve tissues

Comparative transcriptomic analysis between rheumatic (n=32) and non-rheumatic control mitral valves (n=9) revealed extensive gene dysregulated and a pronounce immune-inflammatory signature in RHD. Unsupervised clustering of mRNA expression profiles clearly segregated rheumatic from control samples, indicating distinct disease-associated transcriptional patterns (Figure 1). Differentially expressed genes were predominantly enriched in immune pathways involving B and T lymphocytes, natural killer (NK) cells, monocytes/macrophages, and pro-inflammatory cytokine signalling.

**Figure 1.**
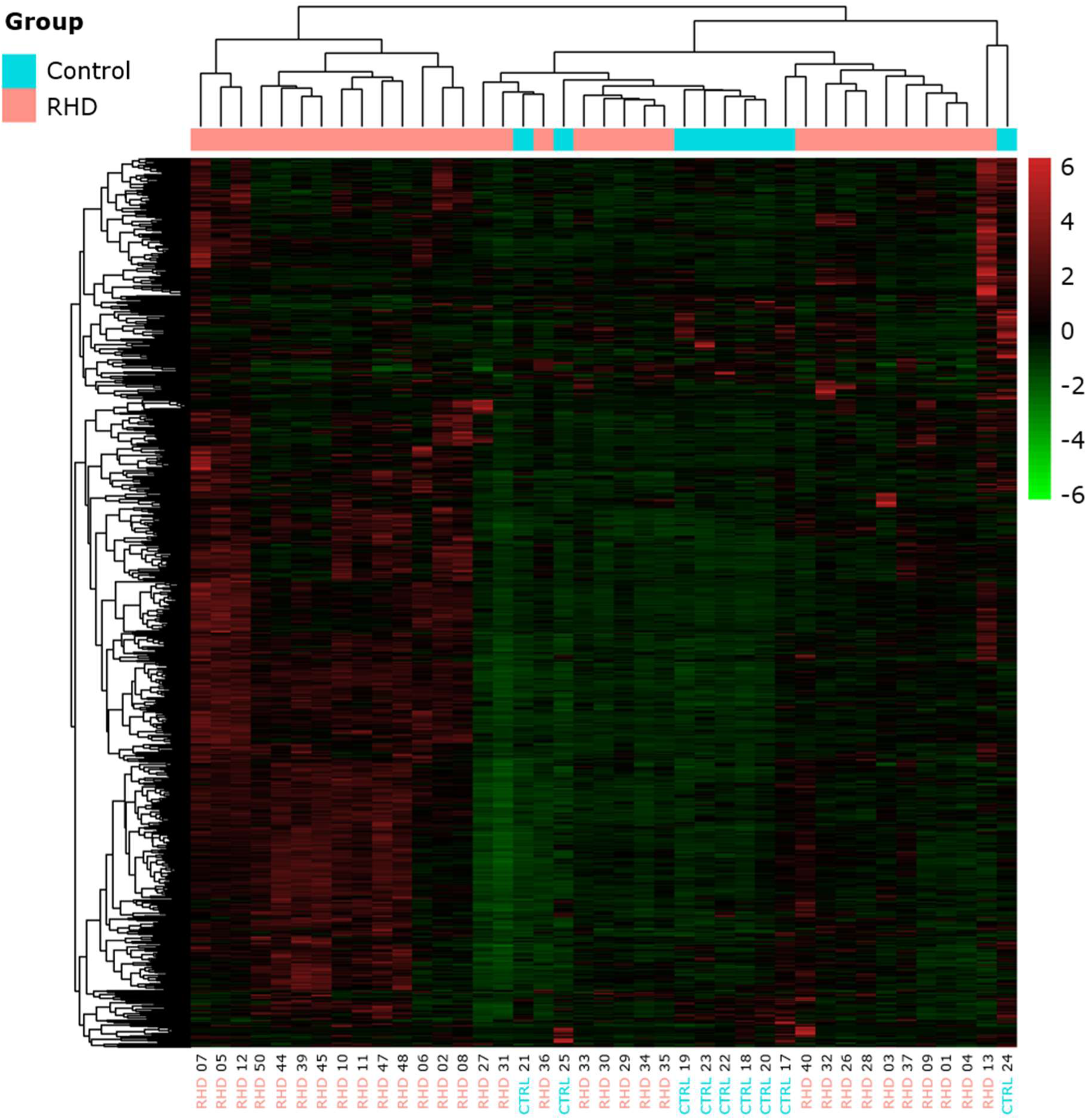
Heatmap of immune-related single gene expression profiles (mRNAs) across rheumatic (RHD) and control (CTRL) mitral valve samples. Each column represents an individual valve sample, and each row corresponds to a single messenger ribonucleic acid (mRNA) included in the analysis. Values are normalized and scaled (z-scores), with red indicating higher relative signal intensity and green indicating lower relative signal intensity, as shown in the colour bar. Unsupervised hierarchical clustering was applied to both samples and mRNAs without prior group labelling. The resulting dendrograms demonstrate a clear structuring of samples into distinct molecular clusters. Notably, samples from patients with RHD predominantly cluster together and are characterized by coordinated upregulation of a large set of immune-related mRNAs, whereas control valve samples show a contrasting pattern with globally lower signal intensity.

Among these immune signatures, B-cell-associated transcripts showed particularly strong and consistent upregulation in rheumatic valves. Differential expression analysis demonstrated marked enrichment of several B-cell markers, including *MS4A1 (CD20)*, *CD19*, *CD22*, *MS4A2,* together with numerous immunoglobulin genes, such as *IGHG1*, *IGHG2*, *IGHG3*, *IGHG4*, *IGHM*, *IGHA1*, *IGKC*, and *IGLC1*, which ranked among the most significantly upregulated transcripts (Figure 2). The coordinated overexpression of multiple independent B-cell markers supports a robust B-cell-associated transcriptional signature in RHD valve tissue.

**Figure 2.**
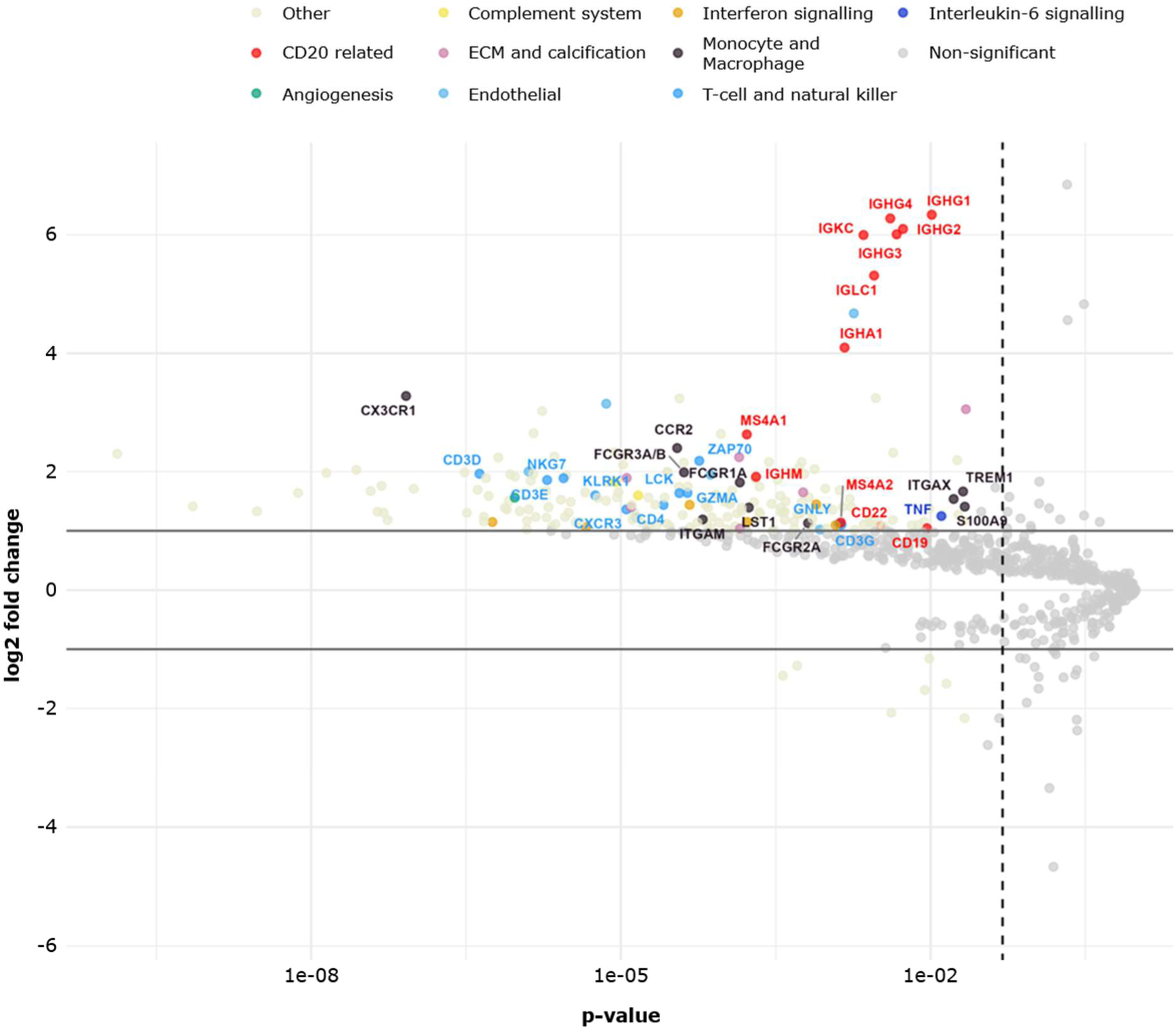
Volcano plot showing differential gene expression between rheumatic (RHD) and control mitral valve tissues. Each dot represents a gene analysed using a predefined immune-related transcriptomic panel. The x-axis shows the p-value (log scale), and the y-axis represents the log2 fold change between RHD and control samples. Genes meeting the significance criteria (p-value < 0.05 and |log2 fold change| > 1) are highlighted and color-coded according to functional immune categories, including CD20-related B-cell genes, T/NK lymphocyte genes, monocyte/macrophage genes, IL-6-related genes, complement genes, and others. The analysis reveals a marked enrichment of significantly upregulated B-cell-associated genes in RHD valves. Notably, immunoglobulin genes (*IGHG1, IGHG2, IGHG3, IGHG4*, *IGHA1*, *IGKC*, *IGLC1*), display both high statistical significance and large effect sizes. These genes are among the most strongly upregulated transcripts in rheumatic valves, indicating increased B-cell presence and/or activation within diseased tissue. In parallel, genes related to T lymphocytes and natural killer cells (*CD3D*, *CD3E*, *CD3G*, *CD4*, *ZAP70*, *KLRK1*, *GNLY*) and to monocyte/macrophage populations (*CCR2*, *CX3CR1*, *FCGR1A*, *FCGR2A*, *FCGR3A*, *ITGAM*, *ITGAX*, *TREM1*, *LST1*, *S100A9*) are also significantly upregulated, reflecting a broader inflammatory infiltrate. Importantly, the clustering of CD20-related and immunoglobulin genes in the upper quadrant of the plot indicates that B-cell-associated pathways represent one of the most prominent and statistically robust differences between RHD and control valves. This finding provides a strong molecular rationale for targeting CD20-positive B-cells in rheumatic heart disease.

Gene-level analyses further confirmed significantly increased expression of *MS4A1*, *MS4A2*, *CD22*, and *CD19* in rheumatic compared with control valves (Figure 3). Similarly, immunoglobulin-related transcripts, including *IGHM*, *IGHA1*, *IGKC*, and *IGLC1*, were markedly increased in RHD samples. Together, these findings are consistent with enhanced B-cell infiltration and/or activation, accompanied by a prominent antibody-producing cell signature within diseased valve tissue.

**Figure 3.**
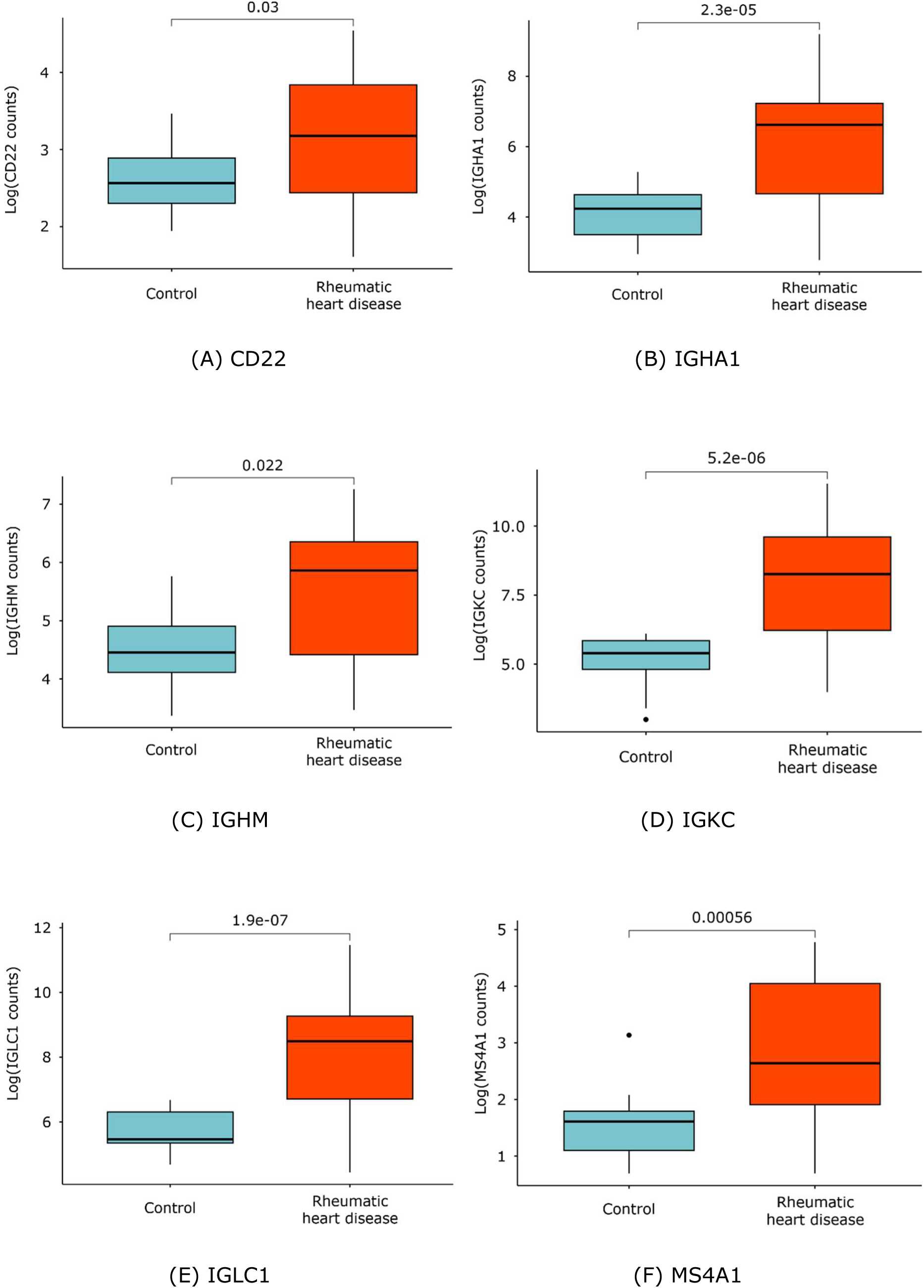
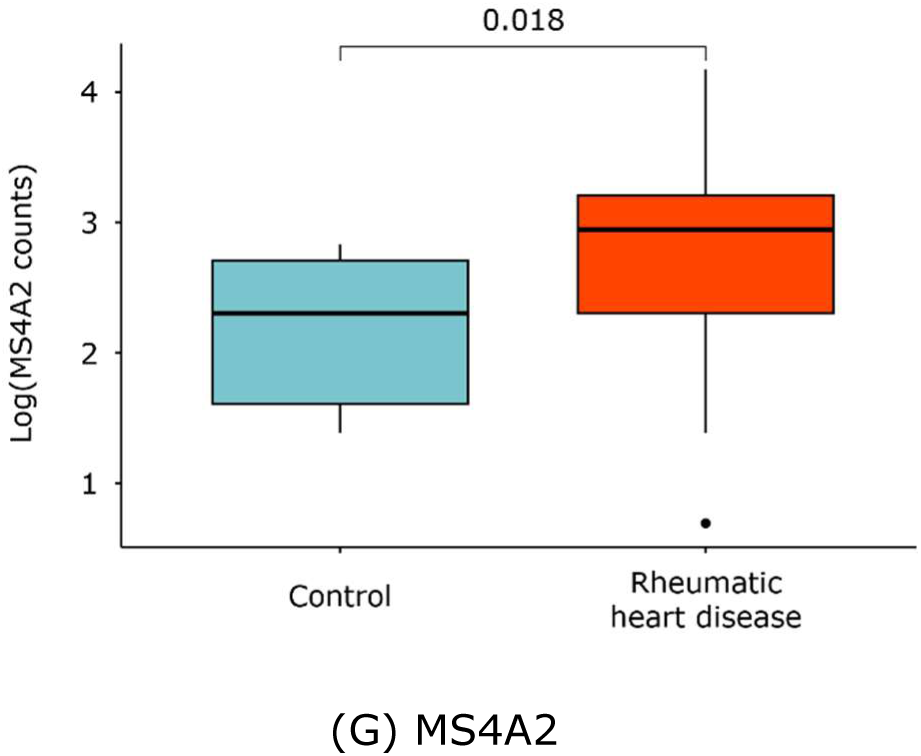
Boxplot representation of selected B-cell-associated and immunoglobulin gene expression levels in rheumatic and control mitral valve tissues. Each panel shows normalized log-transformed gene expression counts for individual genes, comparing control (blue) and rheumatic (red) samples. Statistical comparisons between groups were performed using a Welch t-test, with p-values indicated above each comparison. Genes encoding canonical B-cell surface markers, including *MS4A1* (CD20), *MS4A2*, and *CD22*, show significantly higher expression in RHD valves compared with controls. In parallel, multiple immunoglobulin-related genes (*IGLC1*, *IGHA1*, *IGHM*, *IGKC*) are also markedly upregulated in rheumatic samples, reflecting enhanced local antibody-related activity. The consistent increase across both B-cell identity markers and immunoglobulin genes indicates not only an increased presence of B lymphocytes within rheumatic valve tissue, but also their functional activation. The magnitude and reproducibility of these differences support the conclusion that B-cell-driven immune mechanisms represent a key molecular feature distinguishing rheumatic from control valves.

In agreement with the transcriptomic data, histological analysis demonstrated the presence of CD20-positive B-cells within the inflammatory infiltrate of rheumatic valves (Figure 4). The concordance between transcriptional and histological findings provides complementary evidence for prominent B-cell involvement in the chronic inflammatory response associated with RHD valve pathology.

**Figure 4.**
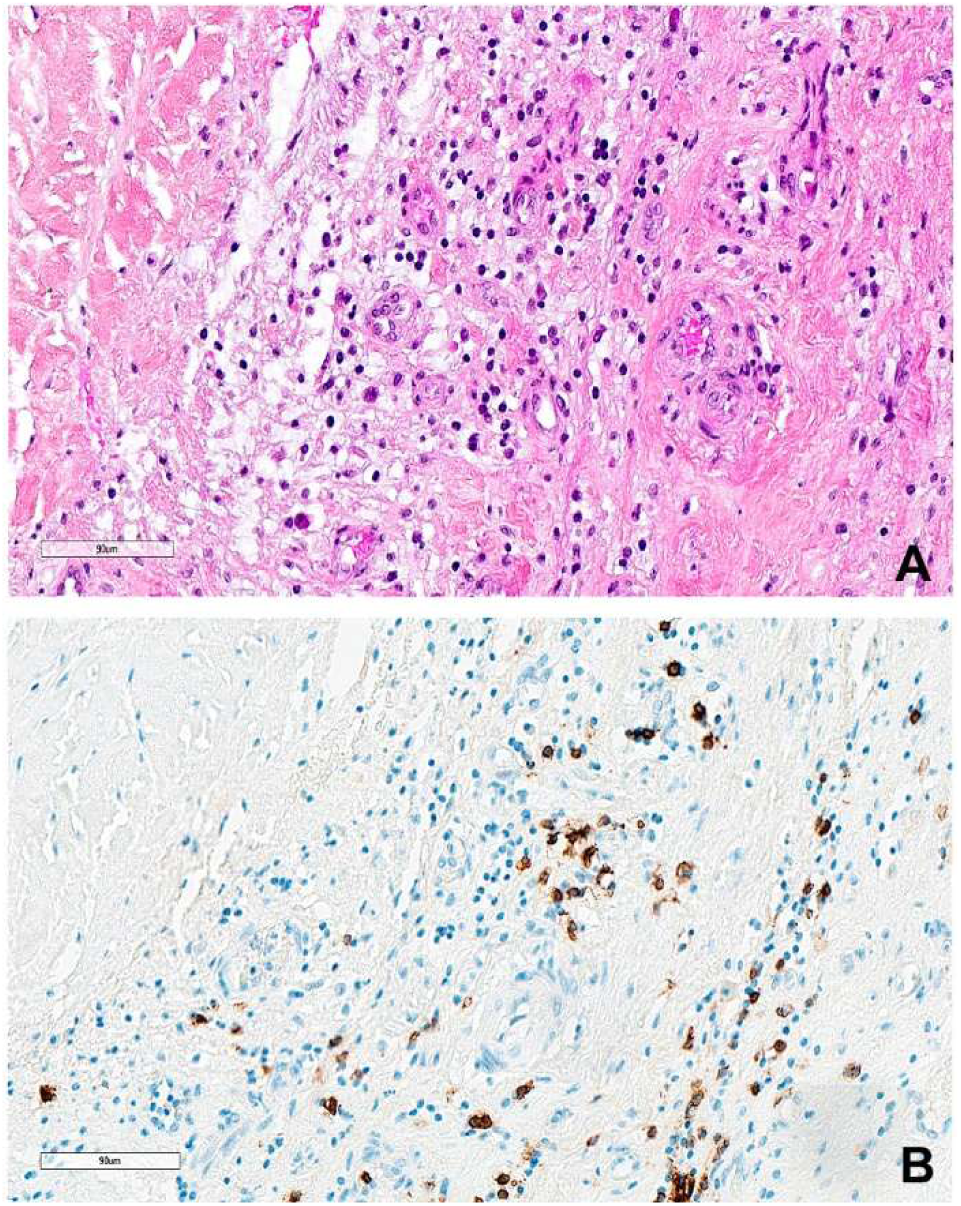
Chronic inflammatory infiltrate with B-cell enrichment in a fibrotic rheumatic aortic valve Adjacent histological sections from the same region of interest in a rheumatic aortic valve, stained with H&E (A) and for CD20 (B). Scale bar, 90 µm. **(A)** Haematoxylin and eosin (H&E) staining showing a dense and heterogeneous inflammatory infiltrate within a fibrous stromal background. The infiltrate is composed of small mononuclear cells with variable morphology, dispersed throughout the tissue and occasionally forming loose aggregates. **(B)** Immunohistochemical staining highlighting the composition of the inflammatory infiltrate. Positive cells are visualized by brown chromogenic staining. CD20 staining reveals scattered B lymphocytes, predominantly distributed in small clusters.

Genes associated with CD4+ helper T cells, CD8+ cytotoxic T cells, and NK cells were also significantly upregulated, including *CD3D*, *CD3E*, *CD3G*, *CD4*, *ZAP70*, *GNLY*, *GZMA*, and *KLRK1*. The increased expression of chemokine receptors such as *CXCR3* suggests active recruitment and retention of these cells within inflamed valve tissue.

In addition, a monocyte/macrophage signature was observed, characterized by elevated expression of *CCR2*, *CX3CR1*, *FCGR1A*, *FCGR2A*, *FCGR3A*, *ITGAM*, *ITGAX*, *TREM1*, *LST1*, and *S100A9*. This profile is consistent with chronic innate immune activation and sustained tissue inflammation.

Key inflammatory mediators were significantly overexpressed in rheumatic valves, notably *TNF*, alongside enrichment of IL-6-related signalling pathways. These findings indicate ongoing inflammatory signalling that may contribute to progressive valvular damage and fibrosis.

Taken together, transcriptomic and expression distribution analyses reveal that rheumatic mitral valves are characterized by a complex immune infiltrate involving B-cells, T/NK lymphocytes, and monocyte/macrophage populations. Based on the particularly strong and reproducible overexpression of CD20-related and immunoglobulin genes, confirmed at both differential expression and distribution levels, we hypothesized that B lymphocytes play a major role in RHD pathophysiology and that B cell depletion using anti-CD20 monoclonal antibody (Rituximab) would be protective. Notably, no suitable animal model currently exists for RHD, further justifying the need for carefully monitored clinical evaluation.

### Feasibility study of anti-CD20 therapy

Having identified CD20-positive B-cells as a plausible therapeutic target, the next step was to evaluate the feasibility and safety of pharmacological B-cell depletion in children with RHD. Rituximab was selected as the anti-CD20 agent because it is a widely available, affordable generic drug in many countries and is well tolerated by paediatric patients.[14] Given the potential risk of impairing humoral immunity in children living in low-resource settings, with inherent increased risk for multiple viral, bacteria and parasite potential infections, we adopted a cautious stepwise approach after approval from the ethical committee.

Since January 2023, fifteen paediatric patients with severe RHD received rituximab in addition to the classical treatment, as part of a feasibility study in the paediatric ward Albert Royer of the Fann University Hospital in Dakar, Senegal. Patients in the intervention group received two infusions, one at baseline (day 0) and a second 14 days later at a dose of 375 mg/m² administered intravenously, to obtain a full and prolonged B cell depletion.[16]

Patients were followed for six months, with particular attention to infusion-related reactions, infections and cardiac adverse events. Two transient complications were observed: vomiting occurred in two patients who had eaten immediately prior to the infusion. After implementing a recommendation that patients should fast before treatment, no further cases were reported. No severe infusion reactions, infectious complications, or delayed adverse events were observed during the six-month follow-up period. Overall, rituximab administration was considered feasible and well tolerated in this preliminary safety cohort. Consequently, and based on this favourable safety profile, it was decided to assess the efficacy of rituximab in a RCT.

### Interim safety analysis

Ethical approval was granted for a RCT (Pan African Clinical Trials registry PACTR202504487625242), contingent upon a planned interim safety analysis after the first year of the study focused on potential side effects (supplementary files).

Two medical doctors (HI, MKS) assessed independently the results. In case of disagreement, the decision was made by a third person blinded for the treatment (EM). Thirty-seven participants (17 in the standard treatment group and 20 in the intervention group), enrolled during the first year, have completed their follow-up and were included in this interim safety analysis. All patients received the standard treatment consisting of oral phenoxymethylpenicillin or erythromycin in cases of severe penicillin allergy. Patients in the intervention group received two infusions of rituximab, one at baseline and a second dose 14 days later. Total Anti-Streptolysin O (ASO) antibody titers, irrespective of immunoglobulin isotype, were measured at baseline before the first infusion, and again 6 months after treatment initiation. ASO antibodies reflect a prior immune response to *Streptococcus pyogenes*, even in the absence of active infection.

In the control group (n=17), the median [IQR] age was 14.0 years [12.0-15.0], with 5 females (29%) and 12 males (71%). In the intervention group (n=20), the median [IQR] age was 11.5 years [10.0-14.2], with 12 females (60%) and 8 males (40%). Two transient complications were observed after anti-CD20 agent infusion. One patient experienced faintness approximately six hours after infusion, associated with a sensation of palpitations. Electrocardiographic (ECG) monitoring did not reveal any abnormal findings. A second patient reported palpitations during the infusion; ECG recording showed a supraventricular arrhythmia, which resolved without further complications. During the 6 months follow-up, two patients required cardiac surgery, one in the intervention group and one in the control group. No death occurred during this 6-month period.

To illustrate the results of the auto immune molecular mechanisms, the clinical and biological results of the first year of the study are given. Among the 32 patients with available ASO measurements, baseline titers were similar between groups. Median [IQR] ASO levels were 400 IU [400–800] in the control group (n=14) and 400 IU [200–700] in the intervention group (n=18), with no statistically significant difference (Mann–Whitney U test, p=0.487). Six months after inclusion, the median ASO titers were 300 IU [200–550] in the control group and 100 IU [100–200] in the intervention group, showing a significant difference between groups (Mann– Whitney U test, p=0.017). A decrease in ASO titers of at least 50% was observed in 12 of 18 patients (66.7%) treated with anti-CD20 agent, compared with 5 of 14 patients (35.7%) in the control group. These results are represented in Figure 5A. Among the 5 participants with missing ASO values (3 in control and 2 in intervention group), the median age was 11 years and 1 (20%) was female.

**Figure 5.**
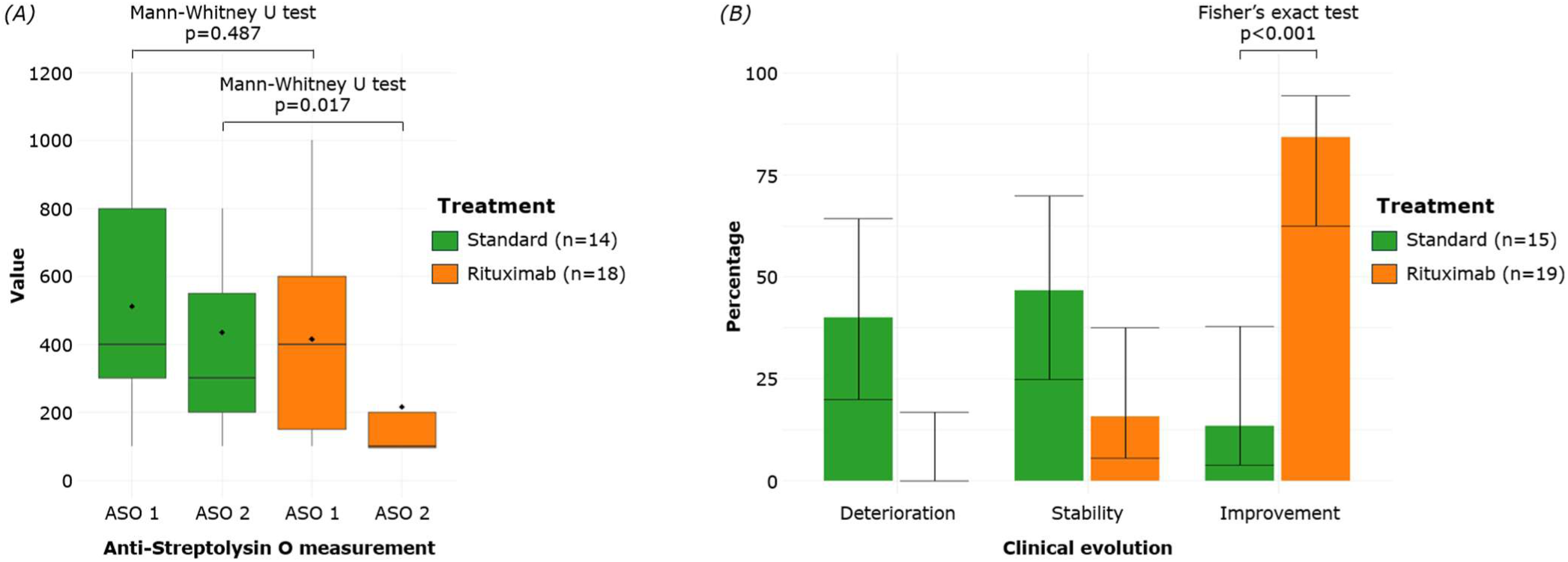
Changes in anti-streptolysin O (ASO) and clinical outcomes according to treatment group with standard treatment on the left and rituximab treatment on the right **(A)** Distribution of ASO levels at baseline (ASO 1) and follow-up values (ASO 2) stratified by treatment group. Boxplots display the median (centre line), interquartile range (box), and whiskers extending to 1.5× the interquartile range; points represent the mean. Among the 32 patients with available ASO measurements, baseline titers were similar between groups. Median [IQR] ASO levels were 400 IU [400-800] in the control group (n=14) and 400 IU [200-700] in the intervention group (n=18), with no statistically significant difference (Mann–Whitney U test, p=0.487). Six months after inclusion, the median ASO titers were 300 IU [200-550] in the control group and 100 IU [100-200] in the intervention group, showing a significant difference between groups (Mann–Whitney U test, p=0.017). **(B)** Distribution of clinical evolution categories (deterioration, stability, and improvement) in patients receiving standard treatment or rituximab. Bars represent the percentage of patients within each treatment group, with error bars indicating 95% confidence intervals estimated using binomial methods. The proportions are calculated relative to the total number of patients in each treatment arm. Clinical outcomes were available for 34 patients. Clinical improvement was assessed on strong criteria (either a return to school, work, or previous physical activities, or a complete disappearance of arthralgia), and was observed in 16 of 19 patients (84.2%) in the intervention group, compared with 2 of 15 patients (13.3%) in the control group. This difference was statistically significant (Fisher’s exact test, p<0.001).

Given these biological modifications, we looked for a potential evolution of the symptoms during this period. Clinical outcomes were available for 34 patients. Clinical improvement was assessed on strong criteria (either a return to school, work, or previous physical activities, or a complete disappearance of arthralgia) and was observed in 16 of 19 patients (84.2%) in the intervention group, compared with 2 of 15 patients (13.3%) in the control group (Figure 5B). This difference was statistically significant (Fisher’s exact test, p<0.001). Among the 3 participants with missing clinical outcomes (2 in control and 1 in intervention group), the median age was 13 years, and all were female.

## Discussion

Our proof of concept provides convergent molecular, histological, biological and clinical evidence supporting a central pathogenic role for B lymphocytes in the pathophysiology of RHD. We show that CD20-positive B cells infiltrate rheumatic valvular lesions and that B-cell depletion with rituximab is feasible and appears relatively safe in children with severe RHD. Together, these findings identify B cells as a potential therapeutic target in a disease for which no disease-modifying treatment currently exists.

RHD remains one of the most neglected cardiovascular diseases worldwide. Although it has virtually disappeared from most high-income countries owing to improved living conditions, widespread antibiotic use and structured prevention programs, it continues to cause substantial morbidity and mortality in low- and middle-income countries, particularly in sub-Saharan Africa.[2],[3] Systematic echocardiographic screening performed 20 years ago by our group and others demonstrated that the burden of childhood RHD was approximately ten-fold higher than previously estimated using World Health Organization case definitions, leading to a reassessment of the global burden of disease.[17] These observations contributed to the revision of the Jones diagnostic criteria in 2015 and to the World Health Organization’s recognition of RHD as a global health priority in 2018.[18],[19]

Despite this renewed attention, treatment strategies remain largely unchanged without any therapeutic innovation. Current management relies almost exclusively on secondary prevention with penicillin, alongside the treatment of ARF and cardiac failures episodes, with valvular surgery considered when necessary, although access remains limited.[3] Although intramuscular benzathine penicillin is highly effective under optimal conditions, its implementation remains challenging because of poor adherence, limited access to healthcare, painful injections and the rare but potentially fatal risk of anaphylaxis.[20] In many countries, intramuscular prophylaxis has progressively been replaced by prolonged daily oral penicillin despite limited evidence regarding adherence and long-term effectiveness. Most importantly, no available immune blocker therapy directly targets the autoimmune mechanisms responsible for progressive valvular injury.[21] Non-specific immuno modulating agents such as Hydroxy Chloroquine have been envisaged to stop the progression from ARF to RHD, and are underway in New Zealand, and different potential targets have been proposed such as Interleukine 6 or TNF alpha, based on their elevated circulatory levels in RHD patients.[22],[23],[24]

RHD has classically been viewed as a post-infectious autoimmune disease initiated by molecular mimicry between Streptococcus pyogenes antigens and cardiac tissues, leading to chronic inflammation, fibrosis and progressive valvular dysfunction.[5],[25],[26] Although pathogenic mechanisms have historically focused on autoreactive T cells, accumulating evidence indicates that B lymphocytes contribute not only through autoantibody production but also through antigen presentation, pro inflammatory cytokine secretion and immune regulation.[13],[23] Our identification of a prominent CD20-centred molecular signature within diseased valves substantially strengthens this concept. Most of the studies have been focused on peripheral blood analyses rather than heart tissues. The rare previous studies that analysed heart valves tissue relied on few candidate-gene approaches generally investigating the expression of several chemokines and their receptors.[27],[28],[29],[30] Our present approach is using the most recent immunologic tools. NanoString profiling enabled the simultaneous analysis of approximately 750 immune-related genes, providing, to our knowledge, the most comprehensive molecular characterization of rheumatic valvular tissue reported to date.

These observations suggest that B cells may play a more sustained role in disease propagation than previously appreciated. Beyond antibody production, they may contribute to chronic valvular inflammation through antigen presentation and secretion of pro-inflammatory cytokines, including IL-6. This interpretation is consistent with observations in other clinical settings, where rituximab-induced B-cell depletion may precede changes in circulating antibody levels. In cryoglobulinaemic vasculitis, clinical improvement may occur before reductions in cryoglobulin or rheumatoid factor levels, while in acute myocardial infarction, rituximab induced rapid B-cell depletion without measurable changes in circulating immunoglobulin levels.[31],[32]

Consistent with this mechanism, rituximab treatment resulted in a marked depletion of circulating B cells together with a significant reduction in ASO titres. Although ASO primarily reflects previous streptococcal exposure rather than ongoing infection, its decline suggests attenuation of persistent humoral immune activation. Rituximab depletes CD20-positive B cells through complement-dependent cytotoxicity, antibody-dependent cellular cytotoxicity and apoptosis while largely preserving T-cell immunity.[33] This selective immunomodulation probably explains the favourable short-term safety profile observed in our cohort despite the high background burden of infectious diseases.[14],[15] Nevertheless, longer follow-up remains essential, particularly to assess immune reconstitution and the potential risk of hepatitis virus reactivation.

Clinical observations should be interpreted cautiously because this proof-of-concept study was not designed to evaluate efficacy. The clinical and biological data presented here primarily serve to support the mechanistic findings. Nevertheless, the rapid improvement observed was accompanied by meaningful functional recovery, including return to school, work or resumption of physical activity. In paediatric populations, these outcomes constitute important indicators of quality of life that may better reflect disease burden than conventional clinical endpoints alone. School attendance is particularly relevant, as prolonged absence may have lasting educational and developmental consequences.

Although preliminary, these findings raise the possibility that B-cell depletion may represent the first disease-modifying therapeutic strategy for RHD. Rituximab has an established role in several autoimmune diseases, including rheumatoid arthritis and systemic lupus erythematosus, and has increasingly become available in resource-limited settings.[34],[35],[36],[37],[38] Its application to RHD has not previously been explored, reflecting the considerable logistical and infrastructural challenges of conducting interventional studies in endemic regions, together with competing public health priorities. In contrast, other targeted immunomodulatory strategies remain largely unexplored, and no alternative biological therapy has demonstrated efficacy in RHD. Preventive vaccination against S. pyogenes represents another promising avenue, with several M-protein and non-M-protein vaccine candidates currently under development; however, no vaccine is yet available for clinical practice.[25],[39],[40],[41],[42],[43],[44],[45]

Several limitations should be acknowledged. First, the clinicians’ awareness of treatment allocation tempers conclusions regarding clinical efficacy and leaves open the possibility of observer and expectation bias. Second, the limited sample size and relatively short follow-up do not allow robust assessment of long-term efficacy, durability of B-cell depletion, immune reconstitution or progression of valvular disease. Third, follow-up in resource-limited settings remains challenging because of socioeconomic constraints, transportation barriers and restricted access to healthcare, resulting in incomplete follow-up for some participants and missing data. In addition, laboratory and imaging resources were necessarily constrained, potentially affecting data completeness and standardization. Finally, as healthcare systems and disease epidemiology vary substantially across sub-Saharan Africa, external validation in other endemic settings will be required before these findings can be generalized. However, these constraints should not be viewed solely as methodological limitations but also as an inherent feature of pragmatic clinical research in endemic settings. If therapeutic innovation for RHD is to benefit the populations most affected by the disease, evidence must be generated under the conditions in which care is actually delivered. Advancing clinical research in low- and middle-income countries therefore requires acknowledging these challenges while investing in the infrastructure needed to progressively overcome them.

An important clinical question is whether B-cell depletion should ultimately be considered earlier in the course of RHD, before irreversible structural valve damage has developed. The current study included only children with advanced valvular disease, and whether earlier intervention could modify the natural history of RHD remains unknown. Of note, three deaths occurred after the first year and were therefore not included in this interim analysis. All deaths were of cardiovascular origin, reflecting the severity of the patients’ underlying condition. Two occurred in the control group and one in the Rituximab group. The ongoing RCT, powered to enrol 108 participants per group, will determine whether rituximab confers durable disease modification rather than transient suppression of inflammation. Echocardiographic assessment of structural valve progression, the key endpoint for establishing disease modification, will require substantially longer follow-up, with definitive data expected in approximately three years. The integration of clinical, biological and echocardiographic outcomes will therefore be essential to establish whether B-cell depletion truly alters disease progression.

This trial also raises an important ethical question. Given the early clinical improvement observed, should treatment continue to be withheld from children randomized to the control group while awaiting structural echocardiographic endpoints? These children remain at risk of prolonged disability, school interruption and progressive cardiac damage. After careful consideration, the Data and Safety Monitoring Board and the ethics committee concluded that continuation of the RCT remained ethically justified because definitive evidence of disease modification has not yet been established.

In conclusion, the concordance between molecular profiling, tissue pathology, biological response and early clinical improvement supports a previously underappreciated role for B cells in RHD pathogenesis. Although causality and long-term disease modification remain to be demonstrated, these findings introduce a mechanistically informed therapeutic strategy for a disease that has seen little innovation for decades. Confirmation in adequately powered randomized trials and in independent endemic populations will determine whether B-cell-targeted therapy can redefine the treatment paradigm for RHD.

## Materials and Methods

### Transcriptomic analysis of rheumatic mitral valve tissues

Mitral valve tissue samples were obtained from 32 patients with severe RHD undergoing surgical valve replacement. Control valve samples were obtained from 9 non-rheumatic valves that included non-immune valvulopathies. All samples were collected at the time of surgery and processed according to local ethical and regulatory approvals.

Gene expression profiling was performed using a nCounter platform (Bruker spatial biology) targeting a predefined panel of 750 mRNAs. This panel was designed to explore inflammatory pathways, immune cell populations, and humoral immune responses, with a particular focus on mechanisms suspected to be involved in autoimmune processes. Raw counts were normalized following the manufacturer’s recommendations. Differential gene expression analysis was conducted to identify significantly dysregulated genes between rheumatic and control valves, using a false discovery rate (FDR) threshold of 0.05 and an absolute log2 fold change of 1.

To support transcriptomic findings, histochemical and histological analyses were performed on valve tissue sections to localize immune cell populations.

### Feasibility study of anti-CD20 therapy

Based on the identification of a strong B-cell signature in rheumatic mitral valves, a study was conducted to assess the safety of anti-CD20 therapy in children with RHD in the paediatric ward Albert Royer of the Fann University Hospital in Dakar, Sénégal.

Rituximab is a chimeric monoclonal antibody of the IgG1 subtype directed against the CD20 antigen, a surface molecule expressed on pre-B and mature B lymphocytes. Binding of rituximab to CD20 leads to B-cell depletion through multiple mechanisms, including complement-dependent cytotoxicity, antibody-dependent cellular cytotoxicity, and induction of apoptosis. Rituximab does not target plasma cells, allowing pre-existing immunoglobulin production to be partially preserved. It is widely used in hematologic malignancies and autoimmune diseases and has a well-characterized safety profile across diverse clinical settings.

Fifteen children with confirmed RHD received rituximab at a dose of 375 mg/m² administered intravenously at baseline (day 0) and a second identical dose 14 days later. Given the potential risks associated with B-cell depletion in children living in settings with high infectious burden, this phase of the study was specifically designed to prioritize safety. Patients were followed for a period of six months after infusion.

Clinical evaluations, biological assessments, and ECG monitoring were performed during scheduled follow-up visits. All adverse events were systematically recorded.

An adverse event was defined as any harmful medical occurrence observed in a participant during the research period, whether or not it was considered related to the investigational product. An adverse effect was defined as any harmful and unintended reaction suspected to be related to the research procedures or to rituximab administration. The attribution of an adverse event as an adverse effect was determined jointly by the investigator and the study sponsor, based on the clinical description, temporal relationship to treatment administration, potential alternative aetiologies, and clinical evolution.

Adverse events were classified as serious if they resulted in death, were life-threatening, required hospitalization or prolongation of hospitalization, caused significant or persistent disability, or resulted in congenital anomaly. Medically important events not meeting these criteria but requiring specific monitoring were also considered serious. Any pregnancy occurring during the study period and any new safety signal potentially modifying the benefit-risk balance were recorded as serious safety events.

The primary objective of this pilot phase was the identification of safety signals associated with rituximab. No efficacy outcomes were evaluated during this phase.

### Interim safety analysis

Then, we proceeded to formally evaluate the potential efficacy of rituximab in an open RCT. Ethical approval was granted, with the requirement to present interim safety analysis after one year focused on potential clinical and biological side effects (Supplementary files). The objective of this analysis was to confirm the safety profile of rituximab during the study. The primary endpoint (echocardiographic rheumatic valvular lesions) was not assessed as part of this interim safety analysis. Therefore, this analysis has no impact on the statistical power, sample size calculation, or planned primary efficacy analysis, all of which will remain unchanged.

The trial is conducted in Senegal at a single tertiary paediatric centre specializing in the management of ARF and RHD. Children aged between 5 and 17 years with confirmed ARF and at least one severe active rheumatic valvular lesion documented by echocardiography were enrolled. Written informed consent was obtained from parents or legal guardians prior to inclusion.

Participants were randomized in a 1:1 ratio to either the intervention group or the control group. Randomization was performed to ensure equal allocation between groups. The absence of blinding was mitigated by the use of objective outcome measures, standardized follow-up procedures, and identical clinical monitoring in both groups.

Inclusion criteria were: confirmed diagnosis of ARF with rheumatic valvular involvement, age between 5 and 17 years, and signed informed consent.

Exclusion criteria included active or chronic infections (HIV, hepatitis B or C, tuberculosis, Epstein–Barr virus), history of severe or recurrent infections, immunodeficiency or autoimmune disease, malignancy, diabetes, recent use of immunosuppressive therapies, prior exposure to monoclonal antibodies, hypersensitivity to rituximab, penicillin, acetylsalicylic acid or paracetamol, recent or planned vaccination, pre-existing heart failure, renal insufficiency (creatinine clearance <45 mL/min/1.73 m²), pregnancy, Guillain–Barré syndrome, abnormal baseline laboratory values (severe cytopenias or elevated liver enzymes), or participation in another interventional trial within the previous six months.

Patients were recruited among children with ARF regularly followed at the paediatric ward Albert Royer of the Fann University Hospital in Dakar, Sénégal. All participants received standard-of-care therapy consisting of oral phenoxymethylpenicillin (1-2 million IU/day in two divided doses for 10 days), or erythromycin in cases of severe penicillin allergy, and acetylsalicylic acid at a dose of 75 mg/kg/day for four weeks.

Patients randomized to the intervention group additionally received rituximab at a dose of 375 mg/m² administered intravenously at baseline (day 0) and a second identical dose 14 days later. Patients in the control group did not receive rituximab but continued standard therapy and additionally received a 5% glucose solution (Glucidion G5) as a placebo, administered intravenously at baseline (day 0) and again 14 days later.

Patients were evaluated at baseline, day 14, and at 3 and 6 months after treatment initiation. Each visit included a clinical examination, blood sampling, ECG, and transthoracic echocardiography.

Biological improvement was defined as a decrease of at least 50% in ASO antibody titers measured 6 months after treatment initiation. ASO titers reflect a prior humoral immune response to *Streptococcus pyogenes* infection, independent of active bacterial presence.

Clinical improvement was defined as either a return to school, work, or previous physical activities, or a transition from arthralgia at inclusion to absence of arthralgia at 6 months, provided that physical activity remained stable and did not deteriorate. Patients were considered stable or worsened if neither of these criteria was met. The table presented in Supplementary file 1 binary encodes the clinical outcomes based on the evolution of arthralgia and activity. Safety outcomes included the occurrence of adverse events, serious adverse events, and new safety signals throughout the follow-up period.

A sample size was calculated for the full RCT; however, for this interim safety analysis, only patients enrolled during the first year of the ongoing study were included. Analyses were conducted on patients with available secondary outcome data.

Baseline ASO titers were compared between the treatment and control groups using the Mann–Whitney U test. ASO titers at 6 months were also compared between the two groups using the Mann–Whitney U test. The proportion of patients exhibiting a clinical improvement was compared between treatment and control groups using Fisher’s exact test. A two-sided p-value < 0.05 was considered statistically significant.

## Data Availability

All data produced in the present work are contained in the manuscript or are available upon reasonable request to the authors.

## List of Supplementary Materials

### Supplementary file 1

Table of binary outcome classification according to the evolution of arthralgia and overall activity status

### Supplementary file 2

Table of ASO and clinical evolution

### Supplementary file 3

Ethical and scientific opinion

### Supplementary file 4

Clinical questionnaire

### Supplementary file 5

Statistical analysis plan

### Supplementary file 6

Histological sections from a rheumatic aortic valve

## Ethics and reporting

This study was conducted in accordance with the Declaration of Helsinki and all applicable ethical guidelines and regulations. Ethical approval was obtained from the relevant ethics committee. Written informed consent was obtained from all participants prior to enrollment, and all relevant documentation was archived in accordance with institutional requirements. The randomized controlled trial was registered in the Pan African Clinical Trials Registry (PACTR) under the registration number PACTR202504487625242.

The authors declare that they have no competing interests.

## Acronyms

ARF: Acute Rheumatic Fever
ASO: Anti-Streptolysin O
CD20: Cluster of Differentiation 20
ECG: Electrocardiogram
FDR: False Discovery Rate
HIV: Human Immunodeficiency Virus
IQR: Interquartile Range
IU: International Unit
mRNAs: Messenger Ribonucleic Acids
NK: Natural Killer
RCT: Randomized Control Trial
RHD: Rheumatic heart disease

## Bibliography

[1] G. A. Roth et al., “Global Burden of Cardiovascular Diseases and Risk Factors, 1990–2019,” JACC, vol. 76, no. 25, pp. 2982–3021, Dec. 2020, doi: 10.1016/j.jacc.2020.11.010.

[2] J. R. Carapetis, A. C. Steer, E. K. Mulholland, and M. Weber, “The global burden of group A streptococcal diseases,” The Lancet Infectious Diseases, vol. 5, no. 11, pp. 685–694, Nov. 2005, doi: 10.1016/S1473-3099(05)70267-X.

[3] E. Marijon, M. Mirabel, D. Celermajer, and X. Jouven, “Rheumatic heart disease,” Lancet, vol. 379, no. 9819, pp. 953–964, Mar. 2012, doi: 10.1016/S0140-6736(11)61171-9.

[4] R. K. Kumar and R. Tandon, “Rheumatic fever & rheumatic heart disease: The last 50 years,” Indian J Med Res, vol. 137, no. 4, pp. 643–658, Apr. 2013.

[5] L. Guilherme and J. Kalil, “Rheumatic Fever and Rheumatic Heart Disease: Cellular Mechanisms Leading Autoimmune Reactivity and Disease,” J Clin Immunol, vol. 30, no. 1, pp. 17–23, Jan. 2010, doi: 10.1007/s10875-009-9332-6.

[6] J. M. Katzenellenbogen et al., “The End Rheumatic Heart Disease in Australia Study of Epidemiology (ERASE) Project: data sources, case ascertainment and cohort profile,” CLEP, vol. 11, pp. 997–1010, Nov. 2019, doi: 10.2147/CLEP.S224621.

[7] A. Beaton, E. Okello, P. Lwabi, C. Mondo, R. McCarter, and C. Sable, “Echocardiography Screening for Rheumatic Heart Disease in Ugandan Schoolchildren,” Circulation, vol. 125, no. 25, pp. 3127–3132, Jun. 2012, doi: 10.1161/CIRCULATIONAHA.112.092312.

[8] L.-A. Gray et al., “Genome-Wide Analysis of Genetic Risk Factors for Rheumatic Heart Disease in Aboriginal Australians Provides Support for Pathogenic Molecular Mimicry,” The Journal of Infectious Diseases, vol. 216, no. 11, pp. 1460–1470, Dec. 2017, doi: 10.1093/infdis/jix497.

[9] A. Beaton et al., “Secondary Antibiotic Prophylaxis for Latent Rheumatic Heart Disease,” N Engl J Med, vol. 386, no. 3, pp. 230–240, Jan. 2022, doi: 10.1056/NEJMoa2102074.

[10] I. Stacey et al., “The evidence that rheumatic heart disease control programs in Australia are making an impact,” Aust N Z J Public Health, vol. 47, no. 4, p. 100071, Aug. 2023, doi: 10.1016/j.anzjph.2023.100071.

[11] K. Hirani et al., “Acute rheumatic fever,” Lancet, vol. 405, no. 10495, pp. 2164–2178, Jun. 2025, doi: 10.1016/S0140-6736(25)00185-0.

[12] S. Marantelli, R. Hand, J. Carapetis, A. Beaton, and R. Wyber, “Severe adverse events following benzathine penicillin G injection for rheumatic heart disease prophylaxis: cardiac compromise more likely than anaphylaxis,” Heart Asia, vol. 11, no. 2, p. e011191, Jun. 2019, doi: 10.1136/heartasia-2019-011191.

[13] G. J. Silverman and D. A. Carson, “Roles of B cells in rheumatoid arthritis,” Arthritis Res Ther, vol. 5, no. S4, pp. S1–S6, Dec. 2003, doi: 10.1186/ar1010.

[14] K. J. Gaffney, E. M. Dahl, M. P. Stanton, E. Starek, and A. S. Zembillas, “Rapid-Infusion Rituximab in a Pediatric Population,” The Journal of Pediatric Pharmacology and Therapeutics, vol. 25, no. 3, pp. 215–219, 2020, doi: 10.5863/1551-6776-25.3.215.

[15] N. J. Olsen and C. M. Stein, “New Drugs for Rheumatoid Arthritis,” N Engl J Med, vol. 350, no. 21, May 2004, doi: 10.1056/NEJMra032906.

[16] J. A. Kari et al., “Rituximab versus cyclophosphamide as first steroid-sparing agent in childhood frequently relapsing and steroid-dependent nephrotic syndrome,” Pediatr Nephrol, vol. 35, no. 8, pp. 1445–1453, Aug. 2020, doi: 10.1007/s00467-020-04570-y.

[17] E. Marijon et al., “Prevalence of Rheumatic Heart Disease Detected by Echocardiographic Screening,” N Engl J Med, vol. 357, no. 5, pp. 470–476, 2007, doi: 10.1056/NEJMoa065085.

[18] M. H. Gewitz et al., “Revision of the Jones Criteria for the Diagnosis of Acute Rheumatic Fever in the Era of Doppler Echocardiography: A Scientific Statement From the American Heart Association,” Circulation, vol. 131, no. 20, pp. 1806–1818, May 2015, doi: 10.1161/CIR.0000000000000205.

[19] World Health Organization, “Executive Board, 141st session, Geneva, 1 June 2017: resolutions and decisions, annexes, summary records,” World Health Organization, EB141/2017/REC/1, 2017.

[20] C. T. Longenecker et al., “Rheumatic Heart Disease Treatment Cascade in Uganda,” Circ: Cardiovascular Quality and Outcomes, vol. 10, no. 11, p. e004037, Nov. 2017, doi: 10.1161/CIRCOUTCOMES.117.004037.

[21] J. R. Carapetis et al., “Acute rheumatic fever and rheumatic heart disease,” Nat Rev Dis Primers, vol. 2, no. 1, p. 15084, Jan. 2016, doi: 10.1038/nrdp.2015.84.

[22] N. Wilson et al., “The roles of immuno-modulator treatment and echocardiographic screening in rheumatic fever and rheumatic heart disease control: research from Aotearoa, New Zealand,” J R Soc N Z, vol. 55, no. 2, pp. 241–266, Apr. 2025, doi: 10.1080/03036758.2024.2306981.

[23] F. M. Middleton, R. McGregor, R. H. Webb, N. J. Wilson, and N. J. Moreland, “Cytokine imbalance in acute rheumatic fever and rheumatic heart disease: Mechanisms and therapeutic implications,” Autoimmunity Reviews, vol. 21, no. 12, p. 103209, Dec. 2022, doi: 10.1016/j.autrev.2022.103209.

[24] R. Webb et al., “Preliminary Cardiac Experience With Hydroxychloroquine in New Zealand Children With Acute Rheumatic Fever,” JAHA, vol. 14, no. 22, p. e043869, Nov. 2025, doi: 10.1161/JAHA.125.043869.

[25] M. W. Cunningham, “Pathogenesis of Group A Streptococcal Infections,” Clinical Microbiology Review, vol. 13, no. 3, pp. 470–511, Jul. 2000, doi: 10.1128/CMR.13.3.470.

[26] M. W. Cunningham, “Rheumatic Fever, Autoimmunity, and Molecular Mimicry: The Streptococcal Connection,” International Reviews of Immunology, vol. 33, no. 4, pp. 314–329, Jul. 2014, doi: 10.3109/08830185.2014.917411.

[27] L. Guilherme et al., “Rheumatic Heart Disease Proinflammatory Cytokines Play a Role in the Progression and Maintenance of Valvular Lesions,” Amercian Journal of Pathlogy, vol. 165, no. 5, pp. 1583–1591, Nov. 2004, doi: 10.1016/s0002-9440(10)63415-3.

[28] Y. Wen, Z. Zeng, C. Gui, L. Li, and W. Li, “Changes in the expression of Th17 cell-associated cytokines in the development of rheumatic heart disease,” Cardiovascular Pathology, vol. 24, no. 6, pp. 382–387, Nov. 2015, doi: 10.1016/j.carpath.2015.07.006.

[29] L. Kim et al., “Overexpression of Transforming Growth Factor-beta1 in the Valvular Fibrosis of Chronic Rheumatic Heart Disease,” J Korean Med Sci, vol. 23, pp. 41–48, Jun. 2007, doi: 10.3346/jkms.2008.23.1.41.

[30] K. Faé et al., “CXCL9/Mig Mediates T cells Recruitment to Valvular Tissue Lesions of Chronic Rheumatic Heart Disease Patients,” Inflammation, vol. 36, no. 4, pp. 800–811, Aug. 2013, doi: 10.1007/s10753-013-9606-2.

[31] P. Cacoub, A. Delluc, D. Saadoun, D. A. Landau, and D. Sene, “Anti-CD20 monoclonal antibody (rituximab) treatment for cryoglobulinemic vasculitis: where do we stand?,” Annals of the Rheumatic Diseases, vol. 67, no. 3, pp. 283–287, Mar. 2008, doi: 10.1136/ard.2006.065565.

[32] T. X. Zhao et al., “Rituximab in patients with acute ST-elevation myocardial infarction: an experimental medicine safety study,” Cardiovascular Research, vol. 118, no. 3, pp. 872–882, Feb. 2022, doi: 10.1093/cvr/cvab113.

[33] J. C. W. Edwards, J. Szechinski, P. Emery, and T. Shaw, “Efficacy of B-Cell–Targeted Therapy with Rituximab in Patients with Rheumatoid Arthritis,” N Engl J Med, vol. 350, no. 25, pp. 2572–2581, Jun. 2004, doi: 10.1056/NEJMoa032534.

[34] E. Kimby, “Tolerability and safety of rituximab (MabThera®),” Cancer Treatment Reviews, vol. 31, no. 6, pp. 456–473, Oct. 2005, doi: 10.1016/j.ctrv.2005.05.007.

[35] J. D. Hainsworth, “Safety of rituximab in the treatment of B cell malignancies: implications for rheumatoid arthritis,” Arthritis Res Ther, vol. 5, no. Suppl 4, p. S12, Dec. 2003, doi: 10.1186/ar1008.

[36] H. Kramm, K. E. Hansen, E. Gowing, and A. Bridges, “Successful Therapy of Rheumatoid Arthritis With Rituximab: Renewed Interest in the Role of B Cells in the Pathogenesis of Rheumatoid Arthritis,” JCR: Journal of Clinical Rheumatology, vol. 10, no. 1, pp. 28–32, Feb. 2004, doi: 10.1097/01.rhu.0000111316.59676.db.

[37] M. J. Leandro, J. C. W. Edwards, and G. Cambridge, “Clinical outcome in 22 patients with rheumatoid arthritis treated with B lymphocyte depletion,” Annals of the Rheumatic Diseases, vol. 61, no. 10, pp. 883–888, Oct. 2002, doi: 10.1136/ard.61.10.883.

[38] J. H. Stone et al., “Rituximab versus Cyclophosphamide for ANCA-Associated Vasculitis,” N Engl J Med, vol. 363, no. 3, pp. 221–232, Jul. 2010, doi: 10.1056/NEJMoa0909905.

[39] D. R. Walkinshaw, M. E. E. Wright, A. E. Mullin, J.-L. Excler, J. H. Kim, and A. C. Steer, “The Streptococcus pyogenes vaccine landscape,” npj Vaccines, vol. 8, no. 1, p. 16, Feb. 2023, doi: 10.1038/s41541-023-00609-x.

[40] A. Fulurija et al., “Research opportunities for the primordial prevention of rheumatic fever and rheumatic heart disease—streptococcal vaccine development: a national heart, lung and blood institute workshop report,” BMJ Glob Health, vol. 8, no. Suppl 9, p. e013534, Dec. 2023, doi: 10.1136/bmjgh-2023-013534.

[41] J. Wang et al., “Streptococcus pyogenes: Pathogenesis and the Current Status of Vaccines,” Vaccines, vol. 11, no. 9, p. 1510, Sep. 2023, doi: 10.3390/vaccines11091510.

[42] É. Pastural et al., “Safety and immunogenicity of a 30-valent M protein-based group a streptococcal vaccine in healthy adult volunteers: A randomized, controlled phase I study,” Vaccine, vol. 38, no. 6, pp. 1384–1392, Feb. 2020, doi: 10.1016/j.vaccine.2019.12.005.

[43] E. Postol et al., “StreptInCor: A Candidate Vaccine Epitope against S. pyogenes Infections Induces Protection in Outbred Mice,” PLoS ONE, vol. 8, no. 4, p. e60969, Apr. 2013, doi: 10.1371/journal.pone.0060969.

[44] L. C. De Sá-Rocha et al., “StreptInCor, a Group A Streptococcal Adsorbed Vaccine: Evaluation of Repeated Intramuscular Dose Toxicity Testing in Rats,” Front. Cardiovasc. Med., vol. 8, p. 643317, May 2021, doi: 10.3389/fcvm.2021.643317.

[45] A. C. Steer, I. Law, L. Matatolu, B. W. Beall, and J. R. Carapetis, “Global emm type distribution of group A streptococci: systematic review and implications for vaccine development,” Lancet Infect Dis, vol. 9, no. 10, pp. 611–616, Oct. 2009, doi: 10.1016/S1473-3099(09)70178-1.

