## Supplemental for "B-cell involvement supports anti-CD20 therapy in paediatric rheumatic heart disease: Proof of concept and initial safety"

#### Title

#### Short title

**Proof of concept for an immune-targeted therapy in paediatric rheumatic heart disease**

#### Authors

Xavier JOUVEN[1,2,3], Magalie KAYA SAMBA[4,5], Hanifa ISMAEL[4,5], Halifa FARCHATI[1,3], Alexandre LOUPY[1,6], Olivier HERMINE[7,8], Marie ANTIGNAC[1,3,9], Patrick BRUNEVAL[1,6], Wolf Herman FRIDMAN[10], Hafid AIT OUFELLA[1], Lamine SY[4,5], Elisabeth TINE[4,5], Ndeye Awa SARR[4,5], Amadou Lamine FALL[5,11], Emma MENANT[1,3], Ibrahima Bara DIOP[4,5]

[1]Université Paris-Cité, Inserm, PARCC, F-75015 Paris, France

[2]Cardiology Department, European Hospital Georges Pompidou, AP-HP, Paris, France

[3]Paris Cardiac Arrest Centre, Paris, France

[4]Cardiology Department, Centre Hospitalier de Fann, Dakar, Senegal

[5]Université Cheikh-Anta-Diop, Dakar, Senegal

[6]Paris Institute for Transplantation and Organ Regeneration, INSERM U970, Université Paris Cité, Paris, France

[7]CEREMAST, Department of Adult Hematology, Necker Hospital, AP-HP, Paris, France

[8]Imagine Institute, INSERM U1163, Université Paris Cité, Paris, France

[9]Department of Pharmacy, Pitié Salpêtrière Hospital, AP-HP, Paris, France
[10]Equipe 13, Centre de Recherche des Cordeliers, INSERM UMRS1138, Université Pierre et Marie Curie/Paris VI, Paris, France
[11]Paediatric Department, Centre Hospitalier de Fann, Dakar, Senegal

### Supplementary Materials

#### Supplementary file 1

| Arthralgia<br>Activity | Deterioration | Stability | Improvement |
|---|---|---|---|
| Deterioration | 0 | 0 | 0 |
| Stability | 0 | 0 | 1 |
| Improvement | 1 | 1 | 1 |

0: Clinical deterioration or stability

1: Clinical improvement

**Table title**

Binary outcome classification according to the evolution of arthralgia and overall activity status

**Table legend**

This table presents a binary encoding of clinical outcomes based on the combined evolution of overall activity status and arthralgia. Rows indicate changes in activity status, while columns indicate changes in arthralgia. Activity improvement corresponds to a return to school, work, or previous physical activities. Arthralgia improvement corresponds to the resolution of arthralgia between inclusion and the 6-month follow-up. Each cell is assigned a value of 0, indicating clinical deterioration or no meaningful improvement, or 1, indicating clinical improvement.

#### Supplementary file 2

| ID | Age range (year) | Sexe | Treatment | ASO 1 | ASO 2 | ASO evolution | Clinical evolution |
|---|---|---|---|---|---|---|---|
| 1 | [10 – 14] | F | Rituximab | 400 | 100 | Decrease | Improvement |
| 2 | [10 – 14] | M | Rituximab | 200 | 100 | Decrease | Improvement |
| 3 | [10 – 14] | F | Rituximab | 400 | 200 | Decrease | NA |
| 4 | [15 – 18] | M | Standard | 100 | 200 | Increase | Stability |
| 5 | [10 – 14] | M | Standard | 100 | 100 | Stabilize | Stability |
| 6 | [10 – 14] | M | Standard | 100 | NA | NA | Stability |
| 7 | [5 – 9] | M | Standard | 300 | NA | NA | Stability |
| 8 | [15 – 18] | M | Rituximab | 100 | NA | NA | Stability |
| 9 | [5 – 9] | M | Rituximab | NA | NA | NA | Improvement |
| 10 | [10 – 14] | F | Rituximab | 800 | 400 | Decrease | Improvement |
| 11 | [10 – 14] | F | Rituximab | 400 | 100 | Decrease | Improvement |
| 12 | [10 – 14] | F | Rituximab | 400 | 100 | Decrease | Improvement |
| 13 | [15 – 18] | M | Standard | 400 | 200 | Decrease | Improvement |
| 14 | [15 – 18] | F | Rituximab | 400 | 100 | Decrease | Improvement |
| 15 | [5 – 9] | F | Rituximab | 1000 | 900 | Stabilize | Stability |
| 16 | [10 – 14] | F | Standard | 400 | 400 | Stabilize | NA |
| 17 | [15 – 18] | F | Standard | 100 | 100 | Stabilize | NA |
| 18 | [10 – 14] | F | Rituximab | 400 | 100 | Decrease | Improvement |
| 19 | [10 – 14] | F | Standard | 400 | 200 | Decrease | Stability |
| 20 | [10 – 14] | F | Rituximab | 200 | 100 | Decrease | Improvement |
| 21 | [15 – 18] | F | Rituximab | 100 | 200 | Increase | Improvement |
| 22 | [15 – 18] | M | Rituximab | 800 | 200 | Decrease | Improvement |
| 23 | [10 – 14] | M | Standard | 400 | 400 | Stabilize | Stability |
| 24 | [10 – 14] | M | Standard | 800 | 200 | Decrease | Deterioration |
| 25 | [10 – 14] | F | Rituximab | 400 | 100 | Decrease | Improvement |
| 26 | [15 – 18] | M | Standard | 400 | 1600 | Increase | Deterioration |
| 27 | [10 – 14] | M | Rituximab | 800 | 800 | Stabilize | Stability |
| 28 | [10 – 14] | M | Standard | 400 | 100 | Decrease | Stability |
| 29 | [10 – 14] | M | Standard | 800 | NA | NA | Deterioration |
| 30 | [15 – 18] | M | Rituximab | 800 | 100 | Decrease | Improvement |
| 31 | [15 – 18] | M | Standard | 800 | 400 | Decrease | Deterioration |
| 32 | [10 – 14] | M | Rituximab | 100 | 100 | Stabilize | Improvement |
| 33 | [10 – 14] | M | Rituximab | 100 | 100 | Stabilize | Improvement |
| 34 | [10 – 14] | F | Standard | 800 | 600 | Stabilize | Stability |
| 35 | [5 – 9] | M | Rituximab | 100 | 100 | Stabilize | Improvement |
| 36 | [15 – 18] | F | Standard | 1200 | 800 | Stabilize | Deterioration |
| 37 | [5 – 9] | M | Standard | 1200 | 800 | Stabilize | Deterioration |

Supplementary file 3

REPUBLIQUE DU SENEGAL
Un Peuple-Un But-Une Foi

Ministère de la Santé
et de l'Action sociale

CNERS Sénégal
La Présidente
Certifié en norme ISO9001-2015

N° 00000051 MSAS/CNERS/SP

Dakar, le 04 MAR. 2025

### AVIS ETHIQUE ET SCIENTIFIQUE

***Référence :*** **Protocole SEN22/20 : « Essai clinique randomisé, contrôlé, en ouvert, évaluant l'efficacité et la tolérance du Rituximab chez des patients présentant un rhumatisme articulaire aigu en Afrique » Version n°3.0 du 18 mai 2024**

**<u>Objet</u>** : Approbation de l'amendement n°1 du SEN22/20 en date du 30 juillet 2024

***Professeur,***

J'accuse réception de vos réponses aux questions relatives au protocole en référence ci-dessus. À l'analyse, le Comité National d'Ethique pour la Recherche en Santé les trouve globalement satisfaisantes. En conséquence, le comité émet un avis éthique et scientifique favorable pour permettre la mise en œuvre dudit protocole.

Cet avis a une durée d'une année à compter de sa date de signature. Son renouvellement reste assujetti à la présentation d'un rapport d'étape permettant d'être informé sur le niveau de mise en œuvre de l'étude. Un rapport de fin de projet vous est exigé pour rester en conformité avec les lois et règlements qui encadrent la recherche pour la santé au Sénégal.

Je vous prie de croire, ***Professeur***, à l'assurance de ma considération distinguée et de mes encouragements renouvelés.

***Pr. Ibrahima Bara DIOP***
***Cardiologue***
***Centre hospitalier universitaire de Fann***
***Dakar, Sénégal***

*English translation*

ETHICAL AND SCIENTIFIC OPINION

Reference: Protocol SEN22/20: "Randomized, controlled, open-label clinical trial evaluating the efficacy and tolerability of Rituximab in patients with rheumatic heart disease in Africa" Version No. 3.0 of May 18, 2024

Subject: Approval of Amendment No. 1 to SEN22/20 dated July 30, 2024

Professor,

I acknowledge receipt of your responses to the questions relating to the above-referenced protocol. Upon review, the National Ethics Committee for Health Research finds them generally satisfactory. Consequently, the committee issues a favorable ethical and scientific opinion to allow the implementation of said protocol.

This opinion is valid for one year from its date of signature. Its renewal remains subject to the submission of a progress report providing information on the level of implementation of the study. A final project report is required of you in order to remain in compliance with the laws and regulations governing health research in Senegal.

Please accept, Professor, the assurance of my highest regard and renewed encouragement.

#### Supplementary file 4

*English translation from French*

Date / /

**CLINICAL QUESTIONNAIRE - AGRAF STUDY - M6 AFTER TREATMENT INITIATION**

**Patient number:** ……………………………………………………………………………………………………

**Date of last treatment administration:** / /

- How do you feel since treatment initiation?

……………………………………………………………………………………………………………………
……………………………………………………………………………………………………………………
……………………………………………………………………………………………………………………

- Did you have joint pain episodes since treatment initiation?

  ☐ Yes ☐ No

If so, how many episodes: ……………………………………………………………………………………

- Did you have fever episodes and/or physical changes (significant weight loss, other)?
  ☐ Yes ☐ No

If so, specify:

……………………………………………………………………………………………………………………
……………………………………………………………………………………………………………………
……………………………………………………………………………………………………………………

- Have you been prescribed corticosteroids (Isone, Cortancyl) when you had these joint pain or fever episodes? ☐ Yes ☐ No

- Do you get short of breath with exertion? ☐ Yes ☐ No

If so, what kind of effort and intensity? Have you noticed an improvement since treatment initiation or not?

……………………………………………………………………………………………………………………
……………………………………………………………………………………………………………………
……………………………………………………………………………………………………………………

- Did you interrupt your usual (standard) treatment at any point?

  ☐ Yes ☐ No

If so, specify the reason and duration of the interruption:

………………………………………………………………………………………………………………………………

………………………………………………………………………………………………………………………………

………………………………………………………………………………………………………………………………

- Have you ever been hospitalized before treatment initiation?

  ☐ Yes ☐ No

If so, specify the frequency:

………………………………………………………………………………………………………………………………

………………………………………………………………………………………………………………………………

………………………………………………………………………………………………………………………………

- Have you been hospitalized since treatment initiation?

  ☐ Yes ☐ No

If so, specify the period and context:

………………………………………………………………………………………………………………………………

………………………………………………………………………………………………………………………………

………………………………………………………………………………………………………………………………

- Did you have to temporarily or permanently interrupt your education because of the disease?

  ☐ Yes ☐ No

If so, is your school attendance more consistent since treatment initiation?

☐ Yes ☐ No

Comments

………………………………………………………………………………………………………………………………

………………………………………………………………………………………………………………………………

………………………………………………………………………………………………………………………………

………………………………………………………………………………………………………………………………

………………………………………………………………………………………………………………………………

………………………………………………………………………………………………………………………………

Supplementary file 5

### Data analyses

**Statistical analysis team**

Statistical analyses will be performed by the team 4 of Inserm Unit U970 under the supervision of Professor Xavier Jouven using validated statistical software, including SAS (SAS Institute Inc., Cary, NC, USA), R (R Foundation for Statistical Computing, Vienna, Austria), and Python.

A detailed Statistical Analysis Plan (SAP) has been finalized before database lock and prior to unblinding of the study data. The SAP prespecifies all analysis populations, endpoint definitions, statistical methods, handling of missing data, subgroup and sensitivity analyses, and the presentation of study results. All statistical analyses will be performed in accordance with the prespecified SAP.

**Sample size calculation**

Assuming an improvement in rheumatic valvular lesions in 40% of participants in the treatment group and 20% of participants in the control group, with a two-sided type I error rate of 5% and 90% power, 108 participants are required in each group, corresponding to a total sample size of 216 participants. Allowing for a 10% rate of loss to follow-up or non-evaluable outcome data, the target sample size will be increased to 120 participants per group, for a total of 240 participants.

**Statistical analysis plan**

*Analysis populations*

The efficacy analysis will be performed in the per-protocol population, defined as participants who complete the 24-month treatment period with satisfactory adherence to the study protocol.

The intention-to-treat (ITT) population, including all randomized participants according to their allocated treatment group irrespective of treatment adherence, will be used for analyses of both efficacy outcomes and safety.

As required by the ethical committee, an interim safety analysis will be conducted after one year of the study, focusing on safety and secondary endpoints. The objective of this analysis is to confirm the safety profile of rituximab during the study. The primary endpoint will not be assessed as part of this interim safety analysis. Therefore, this analysis will have no impact on the statistical power, sample size calculation, or planned primary efficacy analysis, all of which will remain unchanged.

*Descriptive analyses*

Baseline characteristics will be summarized by treatment group. Continuous variables will be reported as mean (standard deviation, SD) for approximately normally distributed data or median (interquartile range [IQR]: 25th–75th percentiles) for non-normally distributed data. Categorical variables will be summarized as counts and percentages.

All statistical tests will be two-sided, and $P < 0.05$ will be considered statistically significant.

*Analysis of the primary and secondary endpoints*

The primary endpoint will be analysed by comparing the proportion of participants with improvement in rheumatic valvular lesions at 24 months between the two treatment groups using Pearson's $\chi^2$ test or Fisher's exact test, as appropriate.

The primary adjusted analysis will be performed using multivariable logistic regression, with improvement in rheumatic valvular lesions at 24 months (yes/no) as the dependent variable. The model will include randomized treatment allocation and baseline disease severity as prespecified covariates. Additional clinically relevant baseline characteristics, identified a priori, may be included to improve the precision of the treatment effect estimate. Treatment effects will be reported as odds ratios (ORs) with corresponding 95% confidence intervals.

Secondary binary endpoints will be analysed using the same approach. Continuous secondary outcomes will be compared using Student's t-test for normally distributed variables or the Mann–Whitney U test for non-normally distributed variables. When comparisons involve more than two groups or repeated measurements, analysis of variance (ANOVA), or the corresponding appropriate model, will be used. Categorical variables will be compared using Pearson's $\chi^2$ test or Fisher's exact test, as appropriate.

*Missing data*

Every effort will be made to ensure complete data collection throughout the study. In particular, ascertainment of the primary endpoint (rheumatic valvular lesion assessment) will be sought for all participants completing follow-up.

For secondary endpoints, the extent and pattern of missing data will be examined. If the proportion of missing data is considered substantial, appropriate multiple imputation methods will be applied under the assumption that data are missing at random. Sensitivity analyses will be performed, where appropriate, to assess the robustness of the findings to different assumptions regarding missing data.

*Protocol deviations and study discontinuation*

Major protocol deviations will include:

- permanent discontinuation of the study treatment;

- inadequate adherence to study procedures;
- treatment allocation errors, which will be identified through reconciliation of the randomization records with the case report forms (CRFs);
- failure to attend scheduled follow-up visits, including loss to follow-up, resulting in missing outcome data;
- unavailable endpoint assessments despite participant attendance (for example, missing echocardiographic evaluation), resulting in missing data.

Participants with major protocol deviations will be excluded from the per-protocol analysis but will remain included in the intention-to-treat analysis whenever possible.

#### Supplementary file 6

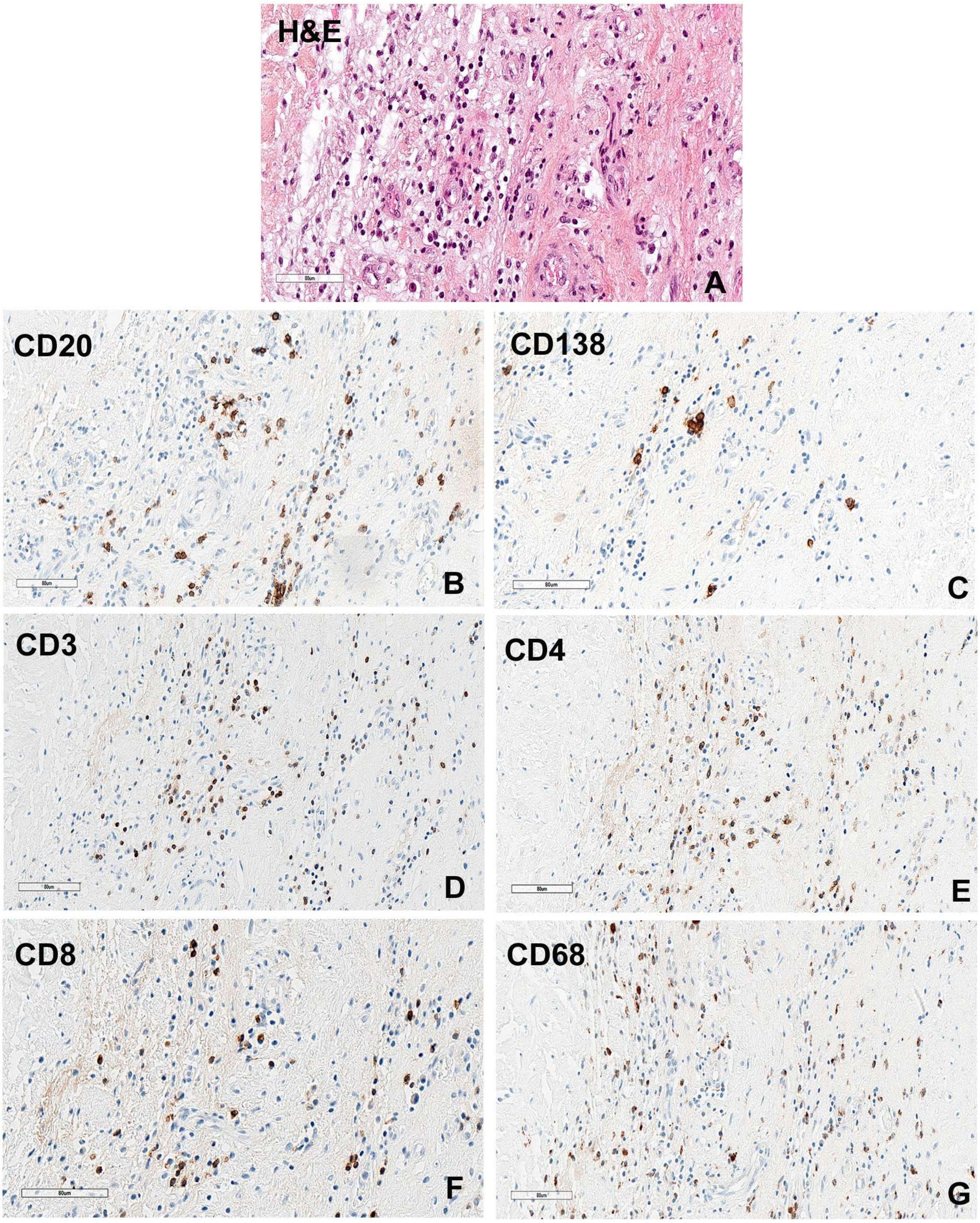


**Figure title**

Chronic inflammatory infiltrate with B-cell enrichment in a fibrotic rheumatic aortic valve

**Figure legend**

Adjacent histological sections from the same region of interest in a rheumatic aortic valve. Scale bar, 90 μm.

**(A)** Haematoxylin and eosin (H&E) staining showing a dense and heterogeneous inflammatory infiltrate within a fibrous stromal background. The infiltrate is composed of small mononuclear cells with variable morphology, dispersed throughout the tissue and occasionally forming loose aggregates.

**(B-G)** Immunohistochemical staining highlighting the composition of the inflammatory infiltrate. Positive cells are visualized by brown chromogenic staining.

(B) CD20 staining reveals scattered B lymphocytes, predominantly distributed in small clusters.

(C) CD138 staining identifies plasma cells, present in lower numbers and dispersed within the infiltrate.

(D) CD3 staining demonstrates a prominent T-cell population throughout the tissue.

(E) CD4 staining highlights helper T lymphocytes, representing a substantial subset of the T-cell infiltrate.

(F) CD8 staining marks cytotoxic T lymphocytes, present in moderate numbers and intermingled with CD4+ cells.

(G) CD68 staining identifies macrophages/histiocytes, diffusely distributed within the stromal component.
